# Segmentation-Guided Development of Visual Classification Criteria for Alzheimer’s Disease

**DOI:** 10.1101/2025.09.29.25336750

**Authors:** Mirjam Peters, David Steinbart, Alexander Hammers, Rolf A. Heckemann, the Alzheimer’s Disease Neuroimaging Initiative

## Abstract

Alzheimer’s disease (AD) causes progressive structural brain changes that precede clinical symptoms by years. Detecting these changes using structural MRI remains challenging, especially in early stages and when relying on visual interpretation alone. Automated semantic segmentation methods offer anatomical precision and objective measurements, but their outputs are rarely used to support human visual assessment.

In this study, we explored whether such segmentation outputs can be used to guide a non-expert investigator in developing and applying interpretable diagnostic criteria. We used images from the Alzheimer’s Disease Neuroimaging Initiative (ADNI) and implemented a structured, segmentation-informed workflow in which a novice with no prior training in radiology or neuroanatomy developed classification rules based on visual appearance and volumetric readouts through three guided pilot phases. In a fourth phase, the investigator applied these criteria to an independent subset of ADNI images while blinded to the respective ADNI participants’ diagnostic labels.

Using an anatomical segmentation model (MAPER) with training data from a pre-release version of the Hammers Adult Brain Atlas Database (120 brain regions), the investigator focused on the piriform cortex (PC). The choice of PC was context-driven, reflecting an ongoing quantitative study of PC volume. A binary classification (AD-like versus CN-like) rule based on PC volume (< or > 430 *mm*^3^), supported by assessments of PC shape and global atrophy, yielded an accuracy of 0.71 across 200 cases spanning four diagnostic groups. Accuracy increased to 0.77 when the analysis was restricted to CN and AD cases (with intermediate pathology (MCI) excluded).

These results show that segmentation-guided visual workflows can enable non-experts to apply anatomically grounded classification criteria with moderate accuracy. Our framework can be expanded to other regions and promises to be useful for generating interpretable models, for supporting explainable AI, and for accelerating the acquisition of diagnostic skills.

## Introduction

Alzheimer’s disease (AD) is a progressive neurodegenerative disorder in which brain structure and function deteriorate gradually, often years before clinical symptoms become evident. Detecting these early structural changes remains a diagnostic challenge. While biomarkers such as amyloid PET and cerebrospinal fluid assays have improved predictive accuracy, magnetic resonance imaging (MRI) remains one of the most widely used and accessible tools for evaluating brain structure in suspected AD cases [1,2]. The primary purpose of structural MRI is currently to exclude alternative, especially reversible, causes of cognitive decline (tumours, vascular lesions, normal-pressure hydrocephalus, and others), rather than to confirm early-stage AD [3].

Visual assessment of MR images relies on expert radiologists examining full image stacks using established criteria. While this process is robust and clinically trusted, it is optimized for detecting gross abnormalities or focal lesions, not for identifying diffuse, subtle atrophy patterns typical of early neurodegenerative disease. Additionally, improvements in spatial resolution increase the stack sizes and visible details, raising cognitive load and interpretive complexity. Even structured rating systems, such as the medial temporal atrophy (MTA) score [4], while reproducible, are limited in specificity and do not consistently detect early-stage disease [3,5].

Automated image analysis, particularly anatomical segmentation, offers a complementary approach (e.g. [6,7]). Such tools extract region-specific measurements with objectivity and reproducibility, and can support individual-level diagnostic reasoning [8–10]. However, segmentation output typically serves as input to statistical classifiers or group-level analyses, rather than being used to enhance visual interpretation. The result is a persistent disconnect: expert visual diagnosis is interpretive but subjective, while automated methods are objective but opaque [11].

We address this gap by proposing a hybrid diagnostic workflow in which automated segmentation using MAPER [12] serves as a scaffold for developing human-interpretable visual criteria. Rather than training a machine to classify cases, we guided a novice investigator (MP) with no prior radiological training through a segmentation-informed process for developing and applying visual classification rules.

We focused on the piriform cortex (PC), a medial temporal lobe structure that is implicated in early AD pathology but is rarely featured in clinical assessments [13,14]. This focus was context-driven, reflecting the availability of the Hammers Adult Brain Atlas Database [15–20] in a pre-release version that included the PC as a distinct anatomical label. In previous work, we showed that MAPER produces accurate segmentations of the PC on the basis of this atlas database, even in cases with known temporal pathology [21]. The investigator refined diagnostic criteria in three pilot phases using images and segmentations from the ADNI2 cohort and then applied them to a test set while blinded to the diagnostic label of the participants in the test set. We report classification performance and examine whether this hybrid workflow enables non-experts to derive anatomically grounded, reproducible diagnostic insight.

Data collection and sharing for this project was funded by the Alzheimer’s Disease Neuroimaging Initiative (ADNI) (National Institutes of Health Grant U01 AG024904) and DOD ADNI (Department of Defense award number W81XWH-12-2-0012).

## Methods

### Study Design

We chose a cross-sectional design using structural MR brain images to develop and test human-interpretable diagnostic criteria for Alzheimer’s disease (AD). The study combined qualitative and quantitative elements and followed a structured, four-phase workflow. The core objective was to evaluate whether segmentation-informed visual criteria could be developed and applied by a novice investigator without prior radiological training (MP, an upper-secondary level student conducting her final-year project). No prior hypotheses were formulated, and the analysis was exploratory.

### Data Source

MR images were obtained from the Alzheimer’s Disease Neuroimaging Initiative Phase 2 (ADNI2) database. All images were T1-weighted structural scans acquired at the screening visit (labelled “ADNI2 Screening MRI-New Pt”) and selected from cognitively normal (CN), early or late mild cognitive impairment (EMCI, LMCI), and AD diagnostic groups (the SMC (subjective memory complaint) was not considered). The EMCI and LMCI groups were pooled and reassigned, based on the most recent diagnostic label provided by ADNI: participants whose label had changed to AD were assigned to the progressive (pMCI) group; all others to the stable (sMCI) group. An initial set of 722 images served as the source pool, from which independent samples were selected for the three pilot phases using stratified randomization without replacement. A final subset of 200 images was selected for classification testing. The sample size was determined pragmatically: based on the time required to analyze cases in the pilot phases, the final cohort size was adjusted to fit within the available time frame for the project.

Information provided to comply with the ADNI Data Use Agreement: The images were obtained from the ADNI database (adni.loni.usc.edu). The ADNI was launched in 2003 as a public-private partnership, led by Principal Investigator Michael W. Weiner, MD. The original goal of ADNI was to test whether serial magnetic resonance imaging (MRI), positron emission tomography (PET), other biological markers, and clinical and neuropsychological assessment can be combined to measure the progression of mild cognitive impairment (MCI) and early Alzheimer’s disease (AD). The current goals include validating biomarkers for clinical trials, improving the generalizability of ADNI data by increasing diversity in the participant cohort, and to provide data concerning the diagnosis and progression of Alzheimer’s disease to the scientific community. For up-to-date information, see adni.loni.usc.edu.

### Image Segmentation and Preprocessing

All MR images were processed using MAPER (Multi-Atlas Propagation with Enhanced Registration), a machine-learning method for automatic anatomical segmentation [12]. MAPER uses prior anatomical labels derived from the Hammers Adult Brain Atlas Database, which is based on manual expert segmentations of high-resolution MR images [, @gousiasAutomaticSegmentationBrain 15 c]. A pre-release version distinguishing 120 regions (plus background) was used. This version included the piriform cortex (PC) as a distinct region. We chose this region to build on our previous work showing PC involvement in AD [21]. Furthermore, we had prevously shown that the neighbouring amygdala is important and involved early [7,22].

Segmentation was performed on the Tetralith high-performance computing cluster at the National Supercomputer Centre, Linköping University, Sweden. Each image was processed with MAPER to produce anatomical label maps. Volumetric estimates were derived and adjusted for intracranial volume as measured with Pincram [23]. Shell scripting was used to open each image in FSLeyes twice, centring the view on the right and left PC, and providing the relevant measures along with the region name and number. The investigator explored the images interactively in FSLeyes, with full access to segmentation overlays, region metadata, and three-plane navigation. As part of her induction, she became familiar with the tools for toggling label-contour map overlays and efficiently navigating the image volume.

Segmentation was used strictly for anatomical guidance and volume estimation. In particular, no artificial neural network or machine-learning model was trained based on the label maps. Further technical details regarding image acquisition, preprocessing, and segmentation parameters have been reported previously [21].

**Figure 1:**
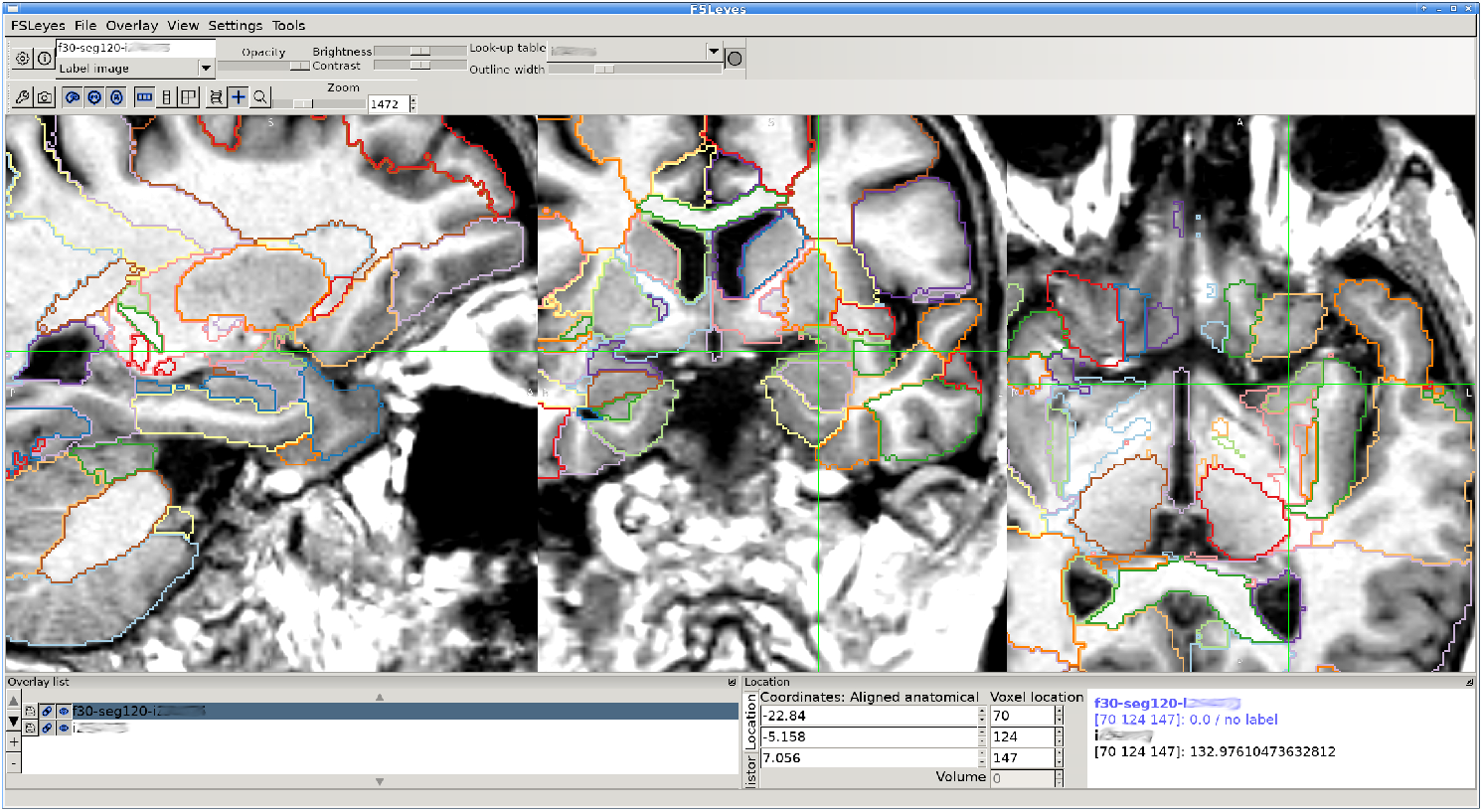
Sample image and label-contour map overlay on a T1-weighted MR image in FSLeyes. The crosshair location is programmatically set to the centre of gravity of one of the PC regions (here: left). The default zoom setting provides a global view; the figure shows a zoomed view of the left medial temporal lobe. Label contours are displayed as hairlines by default; in the figure, the outline width is set to 3 for better visibility.

### Criteria Development Workflow

The development of diagnostic criteria followed three pilot phases, each conducted by the same novice investigator. In the first phase, the investigator examined 14 images (balanced between CN and AD cases) with diagnostic labels disclosed. This initial stage served to familiarise the investigator with the anatomical appearance of cases across the disease spectrum and to identify candidate features for classification. Particular attention was directed toward the piriform cortex and adjacent medial temporal structures, reflecting the common practice of evaluating medial temporal lobe regions as an ensemble.

The design of each subsequent pilot phase was informed by the outcome of the preceding one. Key decisions — including sample size, diagnostic group composition, degree of blinding, and classification objectives — were made by consensus between the investigator and the senior co-author, who served as methodological advisor. An important determinant of this process was the investigator’s self-assessed confidence: the pacing and scope of each pilot phase were adjusted to match her readiness to apply emerging criteria independently. The data used at each stage were removed from the source pool at each stage (sampling without replacement).

During the first pilot phase, the investigator did not acquire the level of confidence she desired. We therefore selected six images (3 CN and 3 AD) to be reviewed in a second pilot phase according to the same approach and priorities as used for the first phase.

In the third pilot phase, 40 images (10 from each diagnostic group: CN, sMCI, pMCI, and AD) were selected. Diagnostic labels were withheld. The investigator was instructed to classify each case as either AD-like or CN-like based on observed patterns. For classification purposes, sMCI was grouped with CN and pMCI with AD, reflecting expected clinical trajectories. The investigator had access to MAPER-derived volumetric data for all regions, but no other metadata. After classification, ADNI-assigned diagnostic labels were disclosed and used to refine a set of visual and quantitative criteria including (1) piriform cortex volume (scalar values separately for left and right, corrected for intracranial volume), (2) PC shape in sagittal and frontal planes as judged visually, and (3) global atrophy as judged visually. The contributions of the criteria were weighted and combined according to the investigator’s subjective impressions.

This subjective weighting turned out to favour the volumetry results to a greater degree than anticipated. Accordingly, we decided to evaluate a purely quantitative classification in addition to the combined approach (cf. next Section “Final Classification Phase”).

### Final Classification Phase

The final test phase involved 200 images (50 per diagnostic group), drawn randomly from the remaining ADNI2 pool, which consisted of 662 images after the pilot samples had been removed (722–14–6–40). The investigator, blinded to the ADNI diagnostic labels but aware of the proportions, applied the refined criteria from the pilot phases and classified each case as either AD-like or CN-like. As in the third pilot phase, sMCI and pMCI were grouped with CN and AD, respectively. For each image, PC volume, PC shape, and overall appearance of atrophy were recorded.

As an additional reference point, we determined diagnostic class predictions (AD-like and CN-like) using the sum volume of the left and right PC, with a cut-off value set to the median of the testing group and treating participants with sub-threshold volumes as AD-like and those with supra-threshold volumes as CN-like. This is reported as the “numeric analysis” in Section Results.

The age distribution in the test set, by diagnostic group, was as follows (min/ median/ max): AD (56.6/77.8/90.4 years), CN (56.3/72.2/84.5), pMCI (55.1/73.6/88.5), and sMCI (56.0/71.4/87.6).

Following classification, diagnostic labels were disclosed. Confusion matrices were generated for two scenarios: the full cohort of 200 cases and a subset containing only the AD and CN groups (n = 100). Performance metrics were calculated, including accuracy, sensitivity, and specificity [24].

## Results

### Classification Performance

In Table 1, four confusion matrices are summarized, based on analysing the full set and the subset; both using numeric analysis and visual analysis. The numeric analysis is based on the median sum volume (left and right piriform cortex) of the group (887 *mm*^3^) as a cut-off.

**Table 1:**
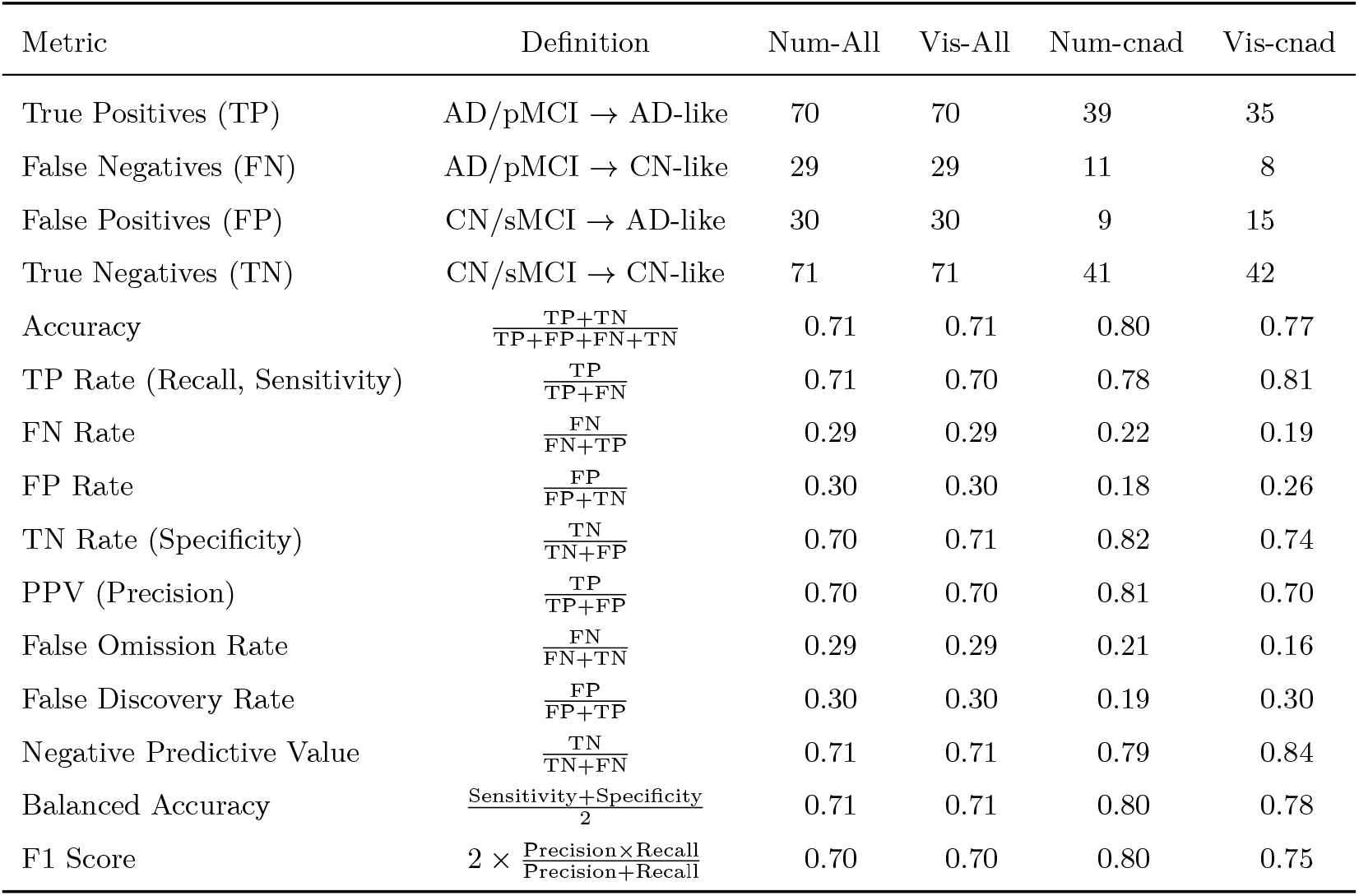
Confusion matrices and performance metrics for numeric (Num) and visual (Vis) analysis of all cases (*n* = 200, “All”) and for CN and AD only (*n* = 100, “cnad”). Read “ →“ as “predicted to be”. PPV: Positive Predictive Value.

- Full Cohort (n = 200) When all diagnostic groups were included, the classification yielded an overall accuracy of 0.71. This level of accuracy is significantly different from chance performance (binomial test; null hypothesis: accuracy 0.5; p = 6 × 10^-9^), indicating that the criteria captured relevant anatomical differences. Key metrics are shown in Table 1.
- CN vs AD Subgroup (n = 100) In the subset containing only CN and AD cases (50 each), the classification accuracy was higher at 0.77 (p = 6 × 10^-8^). This higher performance reflects the greater anatomical separation between unambiguous CN and AD cases. It also underscores the challenge of applying visual criteria to intermediate disease stages.

### Classification Criteria

Three features were identified as useful for classifying cases:

1. Piriform cortex volume: a bilateral average volume threshold of 430 *mm*^3^ (adjusted for total intracranial volume) was used.
  - 430 *mm*^3^: classified as CN
  - < 430 *mm*^3^: classified as AD
2. Piriform cortex morphology
  a. Sagittal View
    - CN: The PC shows a distinct, well-defined morphology. It typically resembles:
      – A horseshoe rotated 90° to the right or
      – A “V” or “Y” rotated 90° to the right
    - AD: The PC appears blurred or amorphous. Some resemblance to the above shapes may persist, but boundaries are less sharp and overall definition is lost.
  b. Frontal View
    - CN: The PC retains a sharply delineated shape. Commonly described as:
      – A horseshoe rotated 90° to the right/left
      – A rotated horseshoe with a longer and narrower caudal arm
    - AD: The PC appears rounded or blob-like, with reduced structural clarity and less distinction from surrounding tissue.
3. Overall Brain Appearance (Global Atrophy)
  - CN (Cognitively Normal): Preserved global brain structure; no clear signs of diffuse atrophy.
  - AD (Alzheimer’s Disease): Evident global atrophy, visible as cortical thinning and ventricular enlargement.

The investigator gained the impression that PC volumes were the most decisive feature. Shape and global atrophy assessments contributed supporting evidence but were insufficient on their own to guide confident classification.

## Discussion

### Principal Findings

A segmentation-guided workflow enabled a novice investigator without prior experience in radiology or neuroanatomy to classify structural MR brain images with moderate accuracy. Using visual criteria developed through structured guidance and informed by automated segmentation, the investigator achieved an overall accuracy of 0.71 across a cohort spanning cognitively normal (CN) participants as well as participants with stable mild cognitive impairment (sMCI), progressive MCI (pMCI), and Alzheimer’s disease (AD). When classification was restricted to CN and AD cases, accuracy increased to 0.76. These results demonstrate that non-experts, given appropriate tools and a structured workflow, can apply anatomically grounded rules with performance well above chance.

### Interpretation

This performance level compares favourably with published interrater agreement for visual diagnosis of AD, particularly given the minimalist rule set, lack of feedback during the classification phase, and absence of prior domain training [3,5]. In fact, radiologists evaluating structural scans often reach only moderate concordance: early studies of medial temporal atrophy (MTA) ratings found interobserver Cohen’s kappa values around 0.6 at best [25]. The strongest predictor in our workflow was a bilateral piriform cortex (PC) volume threshold of 430 *mm*^3^, supported by secondary assessments of PC shape and global atrophy. These simple, interpretable criteria were sufficient to identify many cases accurately, especially at the extremes of the diagnostic spectrum.

The piriform cortex is not commonly included in routine radiological workflows, nor is it standard in public segmentation atlases. Conventional neuroimaging analyses tend to focus on hippocampi, ventricles, and neocortical regions; by contrast, the primary olfactory cortex (of which the PC is a part) is often overlooked or merged with adjacent structures [13,21]. The present project was conducted alongside a quantitative study correlating PC volumes with cognitive test scores (Steinbart D et al., submitted); thus, the focus on PC was context-driven. Whether better classification performance could have been achieved by targeting other regions is not addressed here, and the PC’s neighbouring regions likely influenced the investigator’s impression. Nevertheless, the results underline the value of segmentation tools that provide access to a broad and detailed set of anatomical labels.

Segmentation-informed analysis improves explainability by linking predictions to specific anatomical structures rather than opaque statistical features. Volumetric features such as piriform and hippocampal atrophy are inherently interpretable, and can be verified directly on the images by clinicians. This aligns with current goals in explainable AI, which prioritize transparency and region-level justification of outputs [11].

### Limitations

The classification task was binary: all cases were assigned to either CN or AD, including those with mild cognitive impairment. Grouping sMCI with CN and pMCI with AD was pragmatically motivated by expected clinical trajectories, but inevitably oversimplifies pathological heterogeneity. Diagnostic uncertainty in borderline cases likely contributed to the modest overall accuracy and the observed lower performance when MCI cases were included.

Mild discrepancies in age distributions between diagnostic groups were evident in a manner that reflected the age distribution in the source pool. This could have influenced classification performance. As age is correlated with brain atrophy [26], the investigator may have had an unintended advantage when distinguishing AD from CN cases, as the latter were younger. Future studies should consider explicit age-matching or adjustment of volume thresholds to address this potential bias.

Other limitations include the absence of intra-rater consistency checks or inter-rater validation, and the dependence on a specific segmentation toolchain. Although MAPER provided structural guidance, its performance depends on image quality, registration accuracy, and the chosen atlas. The piriform cortex label used here is not yet part of the standard Hammers atlas release, and reproducibility is therefore contingent on access to an equivalent alternative. Automated volumetric measures are also known to vary across software implementations and atlas definitions [27]. These variations may subtly influence regional estimates and could affect classification outcomes. Further validation using alternate pipelines and segmentation protocols will be needed to confirm generalization.

### Implications and Future Work

These results support the idea that automated segmentation can serve as a scaffold for human visual diagnostics, not merely as an input to statistical or machine learning classifiers. This has implications for explainable AI, where transparency and anatomical interpretability remain major challenges, as well as for radiological education, where guided, region-based workflows may help shorten training curves.

Given the global shortage of radiologists, the results point towards the possibility of engaging less highly trained staff to read images, for example in low-resource settings.

The fact that the numeric classification based on a volume cutoff showed approximately the same accuracy as the combined analysis underlines the value of segmentation as a plausible adjunct to analysis by radiologists. If the stakes are lower than in the typical diagnostic situation, for example when enriching clinical trials with participants who are more likely to progress to AD from MCI, standalone use of automatic segmentation and volumetry can be contemplated.

The approach warrants further investigation. Future studies could explore whether other novice analysts achieve similar performance, whether radiology trainees develop interpretive skills more quickly, whether expert refinement of the criteria yields higher accuracy, and whether the method generalizes to other brain structures or disease contexts. Other possible avenues are to construct a visually-informed severity scale and determine correlations with neuropsychological assessments (e.g. Montreal Cognitive Assessment (“MoCa Test”) available in ADNI2). With appropriate validation, segmentation-informed visual workflows may become a useful component of hybrid diagnostic protocols.

### Conclusion

This study demonstrates that a segmentation-guided workflow can enable non-experts to develop and apply anatomically interpretable classification criteria for Alzheimer’s disease using structural MRI. With minimal prior knowledge, a brief training period, and no feedback during testing, a novice investigator achieved moderate accuracy by combining volumetric and visual features of the piriform cortex. These findings highlight the value of integrating automatic segmentation into visual workflows, both to enhance explainability and to support structured training. Segmentation-informed protocols may help bridge the gap between quantitative image analysis and human interpretability, making them a promising component of future hybrid diagnostic systems.

## Declarations

### Ethics

The ADNI study is conducted under permissions from the respective institutional review boards at the participating medical centres. All participants give written informed consent.

### Author contributions

MP contributed to the study design, developed and tested the classification criteria, calculated the result tables, and wrote a dissertation that served as the basis of the manuscript. DS contributed the manual segmentations of the piriform cortex that are part of the atlas database used for the study, approved the overall study design, and co-edited the manuscript. AH contributed the Hammers Atlas Database, approved the overall study design, and co-edited the manuscript. RAH developed the study concept and design, supervised the pilot studies, carried out the MAPER segmentations, and edited the manuscript.

### Use of generative AI

Portions of this manuscript were developed with the assistance of a large language model (GPT-4o, OpenAI, San Francisco, USA), which was used to support drafting, editing, translation, statistical reasoning, and structuring of the text. The tool did not generate original scientific content or perform any data analysis autonomously. All intellectual contributions, interpretation of results, and final decisions regarding content and conclusions were made by the authors, who take full responsibility for the work.

## Data availability statement

All relevant results are summarized in the manuscript. Individual-level information will not be made available as per the ADNI Data Use Agreement.

The Hammers Adult Brain Atlas Database is available at https://brain-development.org/brain-atlases/adult-brain-atlases/.

